# Advancing Privacy-Aware Machine Learning on Sensitive Data via Edge-Based Continual *µ*-Training for Personalized Large Models

**DOI:** 10.1101/2024.05.18.24307564

**Authors:** Zhaojing Huang, Leping Yu, Luis Fernando Herbozo Contreras, Kamran Eshraghian, Nhan Duy Truong, Armin Nikpour, Omid Kavehei

## Abstract

This paper introduces an innovative method for fine-tuning a larger multi-label model for abnormality detection, utilizing a smaller trainer and advanced knowledge distillation techniques. It delves into the effects of fine-tuning on various abnormalities, noting varied improvements based on the Original Model’s performance in specific tasks. The experimental setup, optimized for on-device inference and fine-tuning with limited computational resources, demonstrates moderate yet promising enhancements in model performance post-fine-tuning. Key insights from the study include the importance of aligning the *µ*-Trainer’s behavior with the Original Model and the influence of hyper-parameters like the batch size on fine-tuning outcomes. The research acknowledges limitations such as the limited exploration of loss functions in multi-label models and constraints in architectural design, suggesting potential avenues for future investigation. While the proposed Naive Continual Fine-tuning Process is in its early stages, it highlights the potential for long-term model personalization. Moreover, using weight transfer exclusively for fine-tuning amplifies user privacy protection through on-device fine-tuning, devoid of transferring data or gradients to the server. Despite modest performance improvements after fine-tuning, these layers represent a small fraction (0.7%) of the total weights in the Original Model and 1.6% in the *µ*-Trainer. This study establishes a foundational framework for advancing personalized model adaptation, on-device inference, and fine-tuning while emphasizing the importance of safeguarding data privacy in model development.

## 1 Introduction

The electrocardiogram (ECG) is a pivotal technology in cardiac dynamics assessment, furnishing insights into the electrical impulses originating from the myocardium. This diagnostic modality holds substantial utility in detecting cardiac irregularities and aberrations, thereby contributing to the early diagnosis of heart abnormalities and subsequent mitigation of stroke risk [1].

Despite the existence of smaller models conducive to operation on edge devices, the incorporation of larger models heralds the prospect of markedly enhanced performance metrics. Such advancements underscore the significance of this approach within the domain of model deployment strategies.

Previously, several studies have focused on compact models, but their performance may not match that of larger models [2, 3, 4, 5]. In this research, we developed a system using knowledge distillation and offsite tuning techniques to fine-tune a large model on an edge device. This approach eliminates the need to transmit user data to the cloud, ensuring patient privacy and saving transmission power. Fig. 1 illustrates the differentiation among conventional methods, methods that share similarities with our proposed approach, and our proposed method.

**Figure 1.**
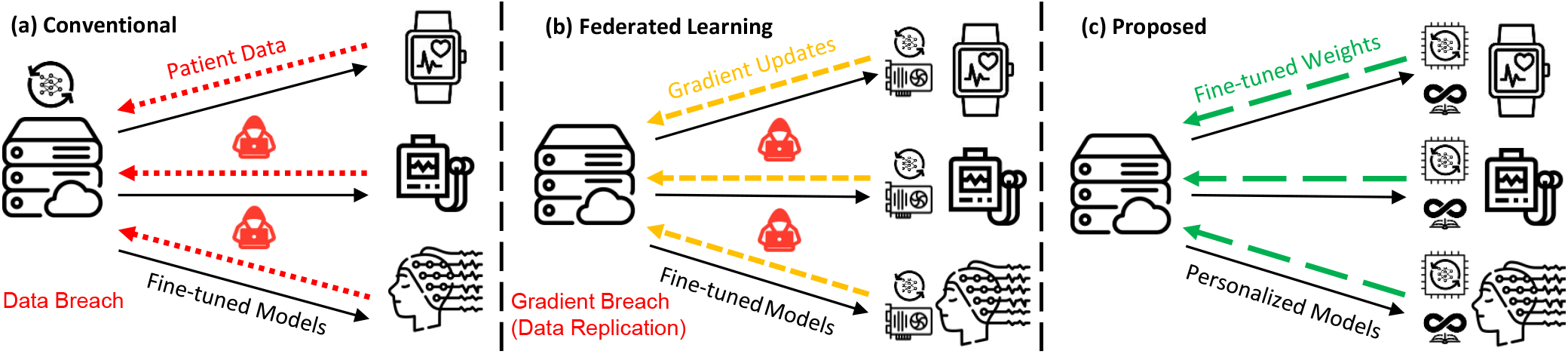
The primary emphasis of this work is on its utilization within wearable devices. a) In contrast to the conventional practice of transmitting data outside the device for fine-tuning, which raises privacy concerns. b) In contrast to Federated Learning, which prioritizes data privacy but necessitates continuous exchange of weight updates during fine-tuning, our approach uniquely addresses the limited fine-tuning capability of devices and enhances model personalization. c) The proposed approach uses a small trainer to fine-tune the data directly on the device. This process updates the original model to generate more personalized inferences, with the potential for continual learning tailored to individual usage patterns.

Privacy and security pose a significant challenge in the AI-driven biomedical field, where balancing data utilization for medical progress with safeguarding individual privacy is paramount. Researchers actively seek solutions to navigate this complex terrain, integrating AI expertise, healthcare knowledge, and legal considerations to address privacy risks effectively [6].

### 1.1 Background

Several models on edge devices have been developed for ECG analysis. Hartmann et al. [7] introduced a Distilled Deep Learning network achieving an accuracy of over 90% for abnormal heartbeat classification. On the other hand, Ram et al. [8] developed a model utilizing random forest for detecting atrial fibrillation, achieving an accuracy of 99%. However, these models may have sub-optimal performance or can only classify a single abnormality. Furthermore, restricting the model’s inference capability to the edge device would impede the device’s capacity to learn and adjust to individual usage patterns. Leveraging large models on edge devices typically necessitates uploading data to the cloud (see Fig. 1(a)), which brings forth several challenges. Privacy becomes a significant concern as transmitting data to the cloud for robust model analysis can result in potential data leaks [9]. Furthermore, the substantial network communication costs are a bottleneck for training models, thereby calling for local edge training solutions [9]. Moreover, extensive data transmission requires a considerable power supply in devices, imposing constraints on device design.

#### 1.1.1 Federated Learning

To address the issue of protecting users’ privacy, a method called Federated Learning (FL) has been developed. This method involves a collaborative and decentralized approach to privacy-preserving technology, aimed at addressing challenges related to data silos and data sensitivity as shown in Fig. 1(b) [10]. FL has been widely adopted in healthcare, including electronic health record management, medical image processing, and remote health monitoring [11].

#### 1.1.2 Federated Learning Vulnerability

However, it still carries the risk of leaking user data. Zhu et al. [12] proposed Deep Leakage from Gradients (DLG), which can recover training inputs and labels from gradients. Boenisch et al. [13] present a method to breach FL protected by Distributed Differential Privacy (DDP) and Secure Aggregation (SA). Both studies demonstrate that FL is not entirely secure for protecting user privacy. In Federated Learning (FL), gradients are calculated using Equ. 1, where *x*_*t*_ and *y*_*t*_ represent a batch of input data and true labels, respectively, and *∇W*_*t*_ denotes the gradient at a specific point in time. *F* () represents the model training within the FL context.

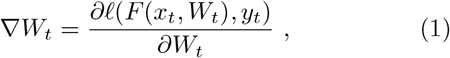

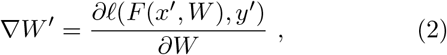

Equ. 2 depicts the process where attackers prepare for data replication, where *x*^*′*^ and *y*^*′*^ are the attacker-constructed dummy and random input data and dummy labels, respectively. The attacker’s knowledge about the data batch can be gained easily due to FL’s training requirements. The attacker constructs dummy input data (*x*^*′*^) and dummy labels (*y*^*′*^) to replicate the data. Using this information, the attacker can match the shape of the original input data and label batch when generating their dummy data and labels. The function *F* () is basically the training model used during the regular operation of FL.

This process allows attackers to achieve the best possible match between the true gradients (*∇W*) and dummy gradients (*∇W*^*′*^), as denoted in Equ. 2, by minimizing the difference. The input data and labels can be replicated through iterative minimization of this gap, as illustrated in Equ. 3. This vulnerability underscores the susceptibility of FL to attacks, as it is shown that the gradients sharing during training can unintentionally create pathways for original data replication and, subsequently, disclosure of sensitive and personal information.

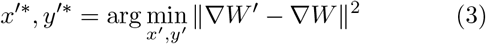

#### 1.1.3 Privacy-Aware *µ*-Trainers

In our study, we’ve introduced a system that allows for on-device fine-tuning of large models, enhancing performance by generating personalized results without the need to transmit data and information externally. To maintain strict access controls over personal data and closely mimic real-world scenarios, we employ a dataset that the model has not previously encountered as a substitute for private personal data. To emulate the very constraint nature of computing power on the edge, we used Raspberry Pi 3 Model B devices to serve as our fine-tuning platform (*µ*-Trainers).

In essence, this work presents a novel approach to addressing on-device training of sizable models while also demonstrating a technique to enhance the privacy and protection of highly sensitive data significantly. In this paper, we chose to show the results of our tests on medical data. We evaluated our proposed technique on two separate datasets of 12-lead electrocardiography (ECG): Telehealth Network of Minas Gerais and China Physiological Signal Challenge 2018 presented in Sec. 3.

We provide an overview of the proposed technique with mathematical explanations in Sec. 1.2, followed by a detailed explanation of the approach in Sec. 4.

### 1.2 Overview of the Proposed Approach

The proposed method, outlined in Fig. 2, is structured into three steps to fine-tune a large model for more accurate personalized usage. Throughout this process, personal data is not transferred beyond the device, mitigating potential privacy concerns. In addition, there is no exchange of information during training, eliminating privacy concerns associated with FL. During the fine-tuning process, information regarding input data batches *x*_*t*_, true labels *y*_*t*_, and gradients *∇W*_*t*_ is kept confidential and not shared beyond the device. Instead, fine-tuning is carried out locally on the device using privately collected data. The iterative batch-level fine-tuning process is described in Equ. 4, and the entire local fine-tuning procedure is outlined in Equ. 5. Subsequently, the fine-tuned trainable weights are shared outside the device.

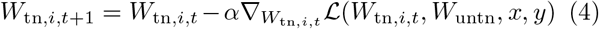

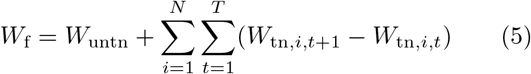

The trainable parameters at iteration *t* + 1 and *t* within epoch *i* are denoted by *W*_tn,*i,t*+1_ and *W*_tn,*i,t*_, respectively. *W*_untn_ represents the fixed, untrainable model weights from the initial pre-trained model. *x* represents the input data, and *y* represents the labels used in the training process. The loss function, denoted as *ℒ*, may depend on the trainable and untrainable weights, input data, and label data. 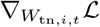 represents the gradient of the loss function with respect to the trainable weights at iteration *t*. The learning rate used in the fine-tuning process is denoted by *α. N* represents the total number of epochs, and *T* represents the total number of iterations within each epoch. *W*_f_ represents the fine-tuned weights of the model.

**Figure 2.**
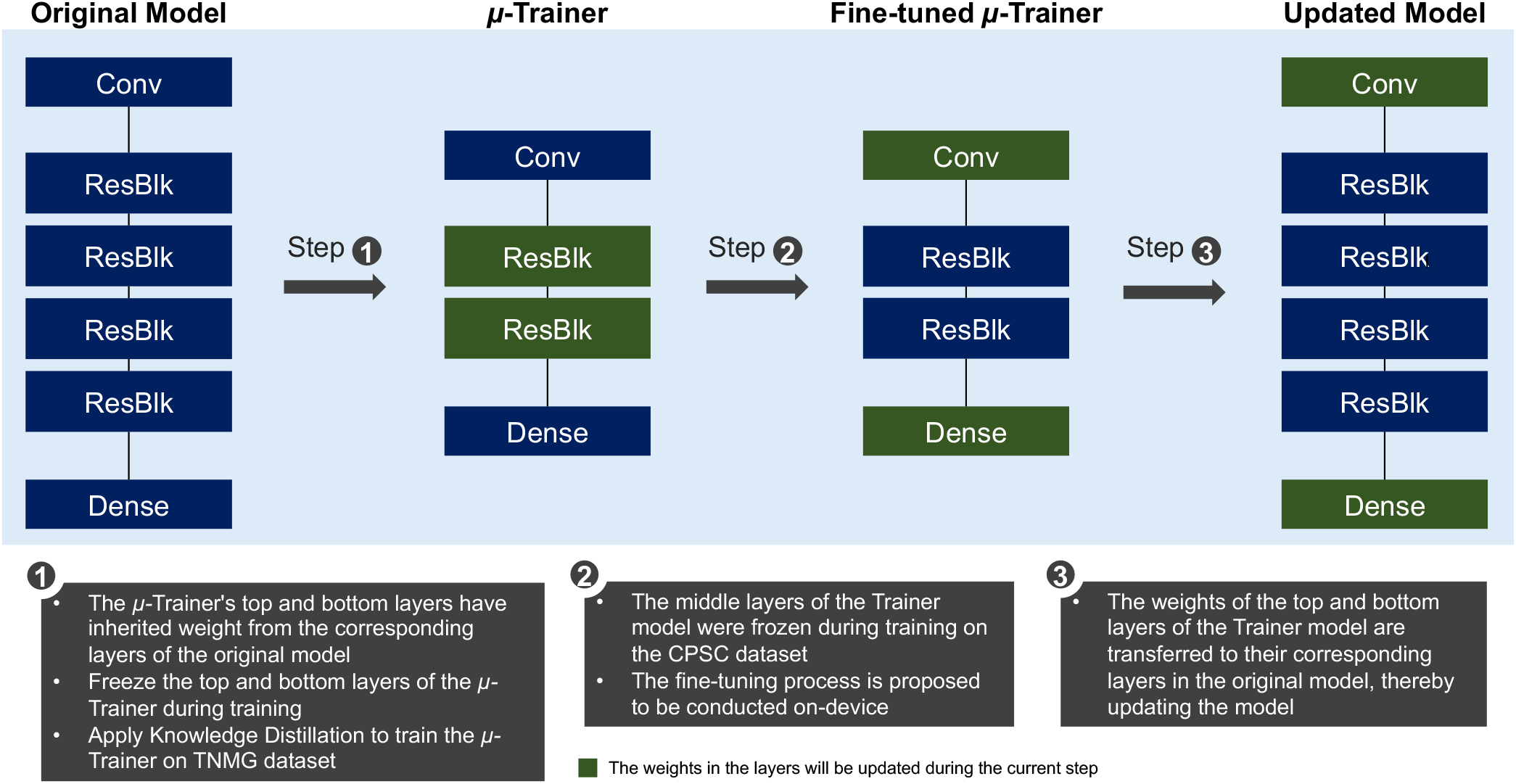
The chart provides a comprehensive overview of the entire method and process, segmented into three distinct steps. In Step 1, weights for the top and bottom layers of the *µ*-Trainer are inherited from the corresponding layers of the original model. These layers are then frozen during training, and Knowledge Distillation is applied to train the *µ*-Trainer on the original dataset. Step 2 involves freezing the middle layers of the Trainer model during fine-tuning on the new dataset on-device. Finally, in Step 3, the weights of the top and bottom layers of the Trainer model are transferred to their corresponding layers in the original model, thereby updating the model. Green highlights in the chart signify that the weights in the layers will be updated during the current step.

A smaller trainer, compact enough to enable on-device training, is utilized to fine-tune the large model, making local training viable. This fine-tuned trainer then updates the weights of the large model, enhancing its ability to provide improved and personalized inferences on the device.

A continuous fine-tuning process, designed to handle incoming data streams over time, was also introduced. This process is outlined in Fig. 3 and extends the previously described method to enable continual fine-tuning.

**Figure 3.**
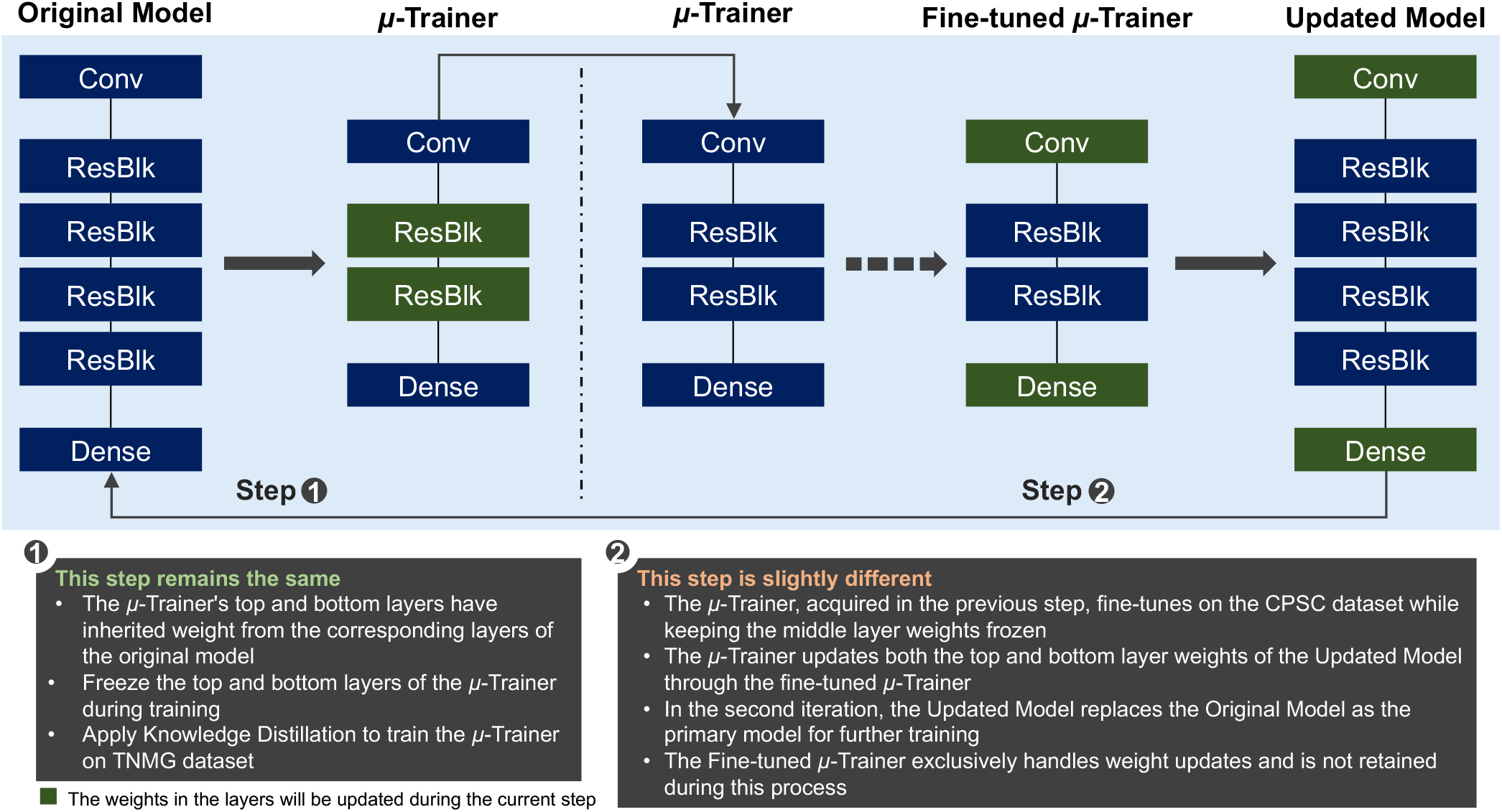
The chart illustrates the proposed continual learning process, which is divided into two distinct steps. In Step 1, the *µ*-Trainer inherits weights for its top and bottom layers from the corresponding layers of the original model, freezes these layers during training, and undergoes Knowledge Distillation to train on the original dataset. Moving to Step 2, the fine-tuned *µ*-Trainer, acquired in the previous step, undergoes further training on the new dataset with its middle layer weights frozen. During this process, the *µ*-Trainer updates the Updated Model’s top and bottom layer weights. Subsequently, in each iteration, the Updated Model replaces the Original Model as the primary model for ongoing training. It’s important to note that the Fine-tuned *µ*-Trainer is exclusively responsible for weight updates and is not retained beyond this process. Layers highlighted in green on the chart signify how weights within those layers are updated.

The detailed approach is further elaborated in the Methods section. Two datasets were utilized to mimic the experiment process while preserving limited personal data. The first dataset represents the original training data for the large model, while the second dataset is used to evaluate the fine-tuning results of the proposed approach. Further details regarding these datasets are provided in the section dedicated to showcasing the datasets.

Compared to other methodologies in this study, our proposed method advances privacy protection, personalization, on-device training, and continuous learning. A qualitative comparison is provided in Table 1 to help better understand the contribution of this work relative to other comparable efforts.

**Table 1:** Comparsion between different methodologies (√; Yes; ×; No; −;Challenging)

|  | Privacy | Personalize | On-device Learning | Continual Learning |
| --- | --- | --- | --- | --- |
| Conventional | $-^\dagger$ | ✓ | $-^\ddagger$ | ✓ |
| FL | × [12] | $-$ [13] | × | ✓ |
| Proposed | ✓ | ✓ | ✓ | ✓ |

$^\dagger$ The conventional method of transmitting user data externally poses significant privacy risks due to the potential for unauthorized access by malicious entities, such as hackers, who can exploit this direct access to sensitive user information.
$^\ddagger$ Edge devices often lack the memory or computational resources needed for the full model training or fine-tuning process, as typically done conventionally.

## 2 Prerequisite

### 2.1 Deep Neural Network

Ribeiro et al. [14] developed a network architecture for ECG detection comprising a sequence of layers. The architecture begins with a convolutional layer (Conv), followed by four residual blocks (ResBlk), each containing two convolutional layers. The final output from the last block is passed through a fully connected layer (Dense) with a Sigmoid activation function *σ*, chosen due to the non-mutually exclusive nature of the classes. Batch normalization (BN) is applied after each convolutional layer, followed by rectified linear activation (ReLU), and dropout regularization is used after the activation. Fig. 4(a) provides detailed insight into the ResNet layer, which forms the basis of the entire model as shown in Fig. 4(b), illustrating the overall model architecture.

**Figure 4.**
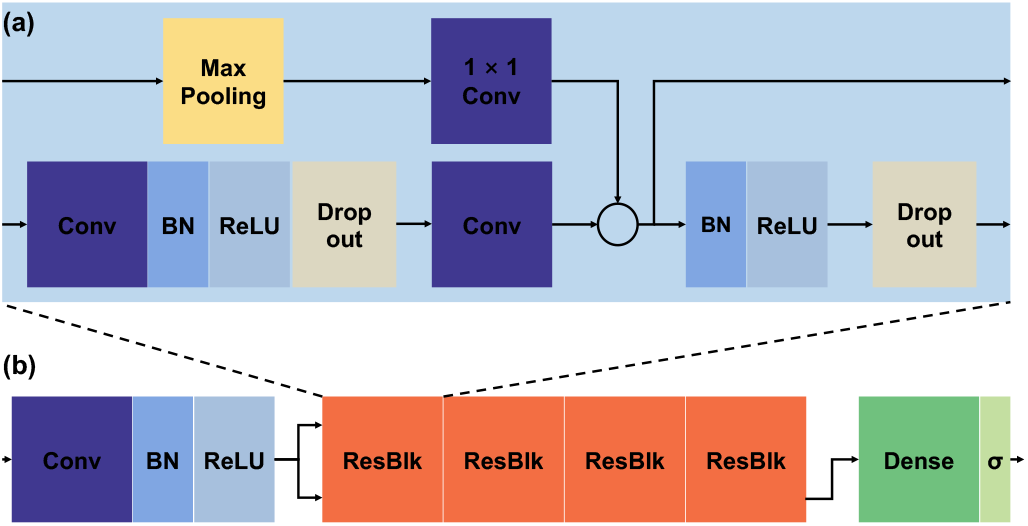
Ribeiro et al. [14]’s model: a) Detailed layout within a ResBlk layer. b) Overview of the model architecture.

The convolutional layers use a filter lengths 16, initially starting with 4096 samples and 64 filters for the first layer and block. The number of filters increases by 64 every second block, and subsampling by a factor of 4 is done in each block. Skip connections with Max Pooling and 1*×*1 convolutional layers are employed to ensure the dimensions match those in the main branch of the network [14]. This model was trained on the Telehealth Network of Minas Gerais dataset.

### 2.2 Knowledge Distillation (KD)

Hinton et al. [15] developed a method that can compress knowledge into a smaller model. The basic theorem behind this is to use a larger model to train a smaller model, which can perform better than training the smaller model directly on the dataset. It is a method that transfers knowledge from a cumbersome model to a smaller model, making it more suitable for deployment.

One effective strategy for transferring the generalization ability of a complex model to a smaller one is to utilize the class probabilities generated by the complex model as “soft targets” during the training of the smaller model. This transfer process can involve either using the same training dataset or a distinct “transfer” dataset [15]. In cases where the complex model consists of a large ensemble of simpler models, we can compute the arithmetic or geometric mean of their predictive distributions to form the soft targets.

Soft targets with high entropy offer significantly more information per training instance than challenging targets. Moreover, they exhibit lower variance in gradient across training instances. Consequently, the small model can often be trained with substantially less data than the original complex model, allowing for a higher learning rate [15].

Leveraging soft targets derived from the complex model enhances the transfer of generalization capabilities to the smaller model, facilitating more efficient training with reduced data requirements and higher learning rates. The resulting model from this method can exhibit greater accuracy and behavior similarity to the large model compared to a model trained directly on the dataset.

### 2.3 Offsite-Tuning

To strengthen the capacity for deploying substantial models on edge computing platforms, Xiao et al. [16] have introduced a methodology. This approach entails the development of an adapter, which undergoes training by a large model and subsequent fine-tuning on an edge device. Notably, the adapter’s refined weights are relayed back to the large model, facilitating a seamless integration between the large model and the edge computing infrastructure.

The original model architecture, *M* = [*A, ϵ*], comprises a trainable adapter *A* and the remaining portion *ϵ* of the model. A lossy comparison is conducted with the emulator component *ϵ*, resulting in a compressed representation *ϵ*^*∗*^. Consequently, the new model is [*A, ϵ*^*∗*^], where fine-tuning updates adapter *A* to *A*^*′*^. The updated adapter *A*^*′*^ is integrated back into the original model, forming *M*^*′*^ = [*A*^*′*^, *ϵ*]. This iterative process enhances model performance on fine-tuned datasets while maintaining similarity to the original frozen component. This approach confines data locally, improving privacy protection [16].

To improve performance across tasks, a sandwich design *M* = *A*_1_ *◦ϵ◦A*_2_ is used, employing both shallow and deep layers, where *A*_1_ and *A*_2_ are the top and bottom layers of the original model and are going to be fine-tuned [16]. The emulator component is intentionally more minor than the original model, enhancing computational efficiency and adaptability. A layer-drop-based compression method is considered, where a subset of layers is uniformly dropped from the original model, and the remaining part is used as the emulator. Notably, retaining the first and last layers is beneficial in this approach [16]. The emulator is further trained with KD under the supervision of the original component *ϵ* with the pre-training dataset and mean squared error as shown in Equ. 6 as the loss function, where *x*_*i*_ refers to the hidden representation of the *i*-th input sample produced by previous layers *A*_1_, *N* refers to the number of samples in the pre-training dataset.

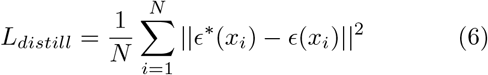

After comprehensively evaluating the prerequisites, it is enlightening to explore a system that enables on-device fine-tuning for improved personalized inference results. This approach maintains data privacy and eliminates the need to transfer large amounts of data outside the device.

## 3 Datasets

This study utilized two distinct datasets to evaluate the proposed models. The first dataset, known as the Tele-health Network of Minas Gerais (TNMG) dataset [14], is specifically designed for automatically detecting abnormalities in rhythm and morphology within 12-lead ECGs. The Original Model used in the study was initially trained on this dataset.

The second dataset utilized in this research is the CPSC dataset (12-lead ECG dataset), developed for The China Physiological Signal Challenge 2018 [17]. This dataset contains abnormality labels for 12-lead ECG data and was explicitly employed for the fine-tuning process to simulate scenarios involving new and unseen data reception.

### 3.1 TNMG

The full TNMG dataset consists of 2,322,513 uniquely different labeled samples, each representing 10 seconds of 12-lead ECG data. These samples cover six distinct types of abnormalities: Atrial Fibrillation (AF), First Degree Atrioventricular Block (1dAVb), Left Bundle Branch Block (LBBB), Right Bundle Branch Block (RBBB), Sinus Bradycardia (SB), and Sinus Tachycardia (ST) [14]. Initially, the ECG data was sampled at a frequency of 400 Hz.

For model training, a balanced dataset was created by randomly selecting 3000 data points for each of the six abnormalities and an additional 3000 data points without any abnormalities, resulting in 21,000 samples. In cases where patients exhibited multiple abnormalities, any remaining samples needed to reach the subset size of 21,000 were chosen randomly from the TNMG dataset. Refer to Table 2 for detailed information about these six abnormalities.

**Table 2:**
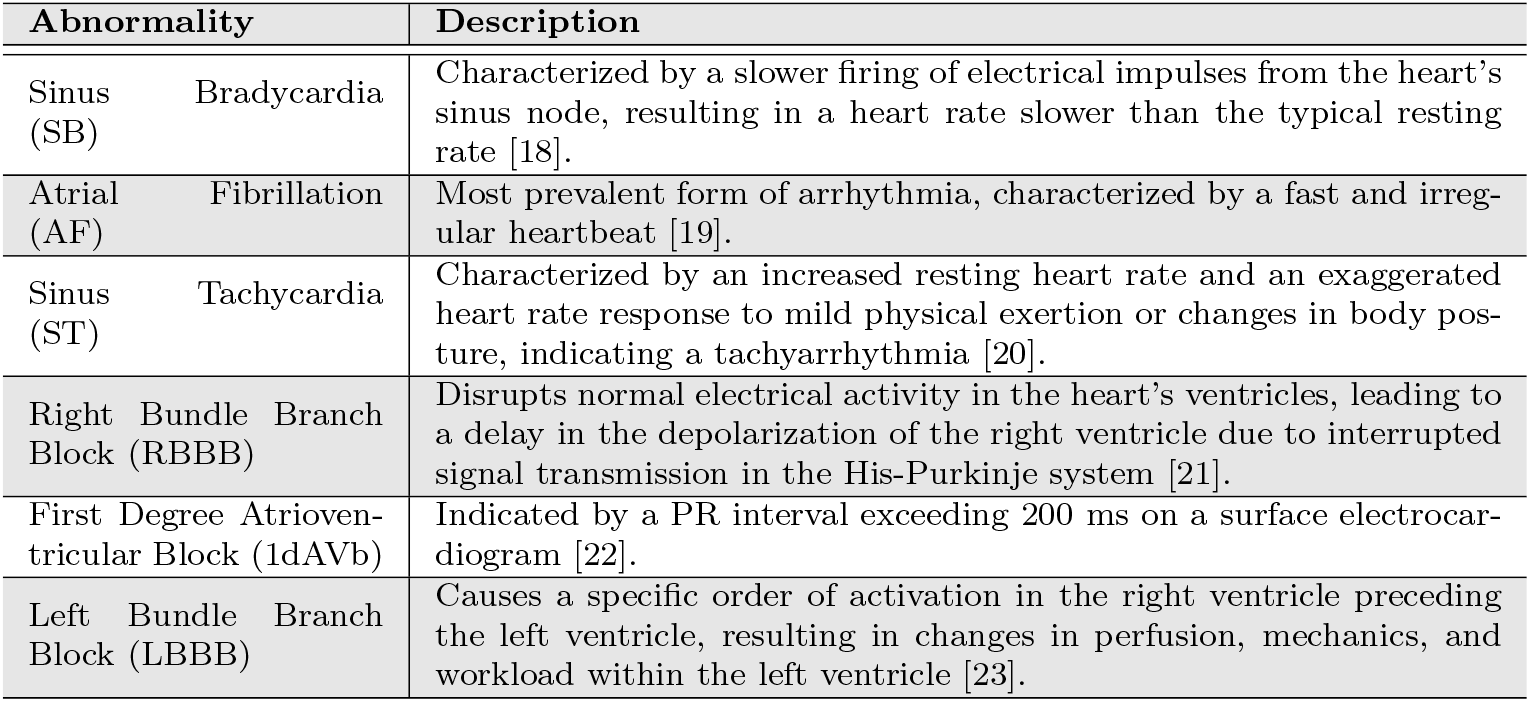
Abnormalities within the TNMG Dataset Categorized into Different Classifications.

The dataset underwent normalization to a consistent length of 4096 readings, ensuring uniformity for analysis and modeling purposes. Readings exceeding this length were removed to streamline data processing and comparison. Fig. 5 illustrates a balanced distribution of genders in the resampled dataset, promoting inclusivity and valid analysis. The dataset also reflects the age distribution observed in the general population, enhancing representativeness for age-related analysis. Additionally, the balanced distribution of different abnormalities in the dataset improves the model’s learning process and overall performance [24].

**Figure 5.**
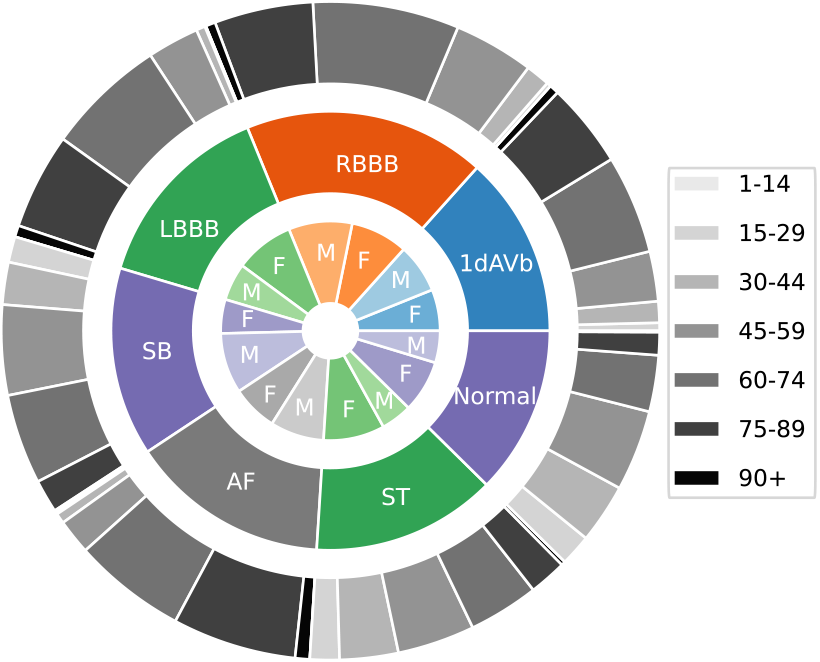
The TNMG subset represents a highly balanced dataset containing six types of abnormalities and exhibits a higher concentration of older patients, reflective of the general population.

### 3.2 CPSC

The CPSC dataset consists of 12-lead ECGs ranging from 6 to 60 seconds, each recorded at a sample rate of 500 Hz. For compatibility with the CPSC dataset, the TNMG data was resampled at a rate of 500 Hz specifically for training purposes. This dataset includes ECGs from patients diagnosed with various cardiovascular conditions and exhibiting common rhythms, with accurate annotations for abnormalities. Overall, the dataset encompasses eight distinct types of abnormalities.

The CPSC dataset, which is new and unseen by the Original Model, is employed to assess the fine-tuning potential of the proposed methods. Despite containing eight (8) classes of abnormalities, four (4) of these classes overlap with those in the TNMG dataset, making them the key classes used to evaluate the proposed fine-tuning strategy. The four chosen abnormalities for evaluation are AF, LBBB, RBBB, and 1dAVb. It’s important to note that this study does not include four other types of abnormalities: Premature Atrial Contraction (PAC), Premature Ventricular Contraction (PVC), ST-segment Depression (STD), and ST-segment Elevated (STE).

As part of the data selection process, entries with missing readings were excluded, resulting in a final dataset of 6,877 distinct ECG tracings. The data was standardized to 4,096 readings for analysis purposes, with any additional readings removed during data cleaning. Fig. 6 provides detailed insights into the distribution of abnormalities within the CPSC dataset.

**Figure 6.**
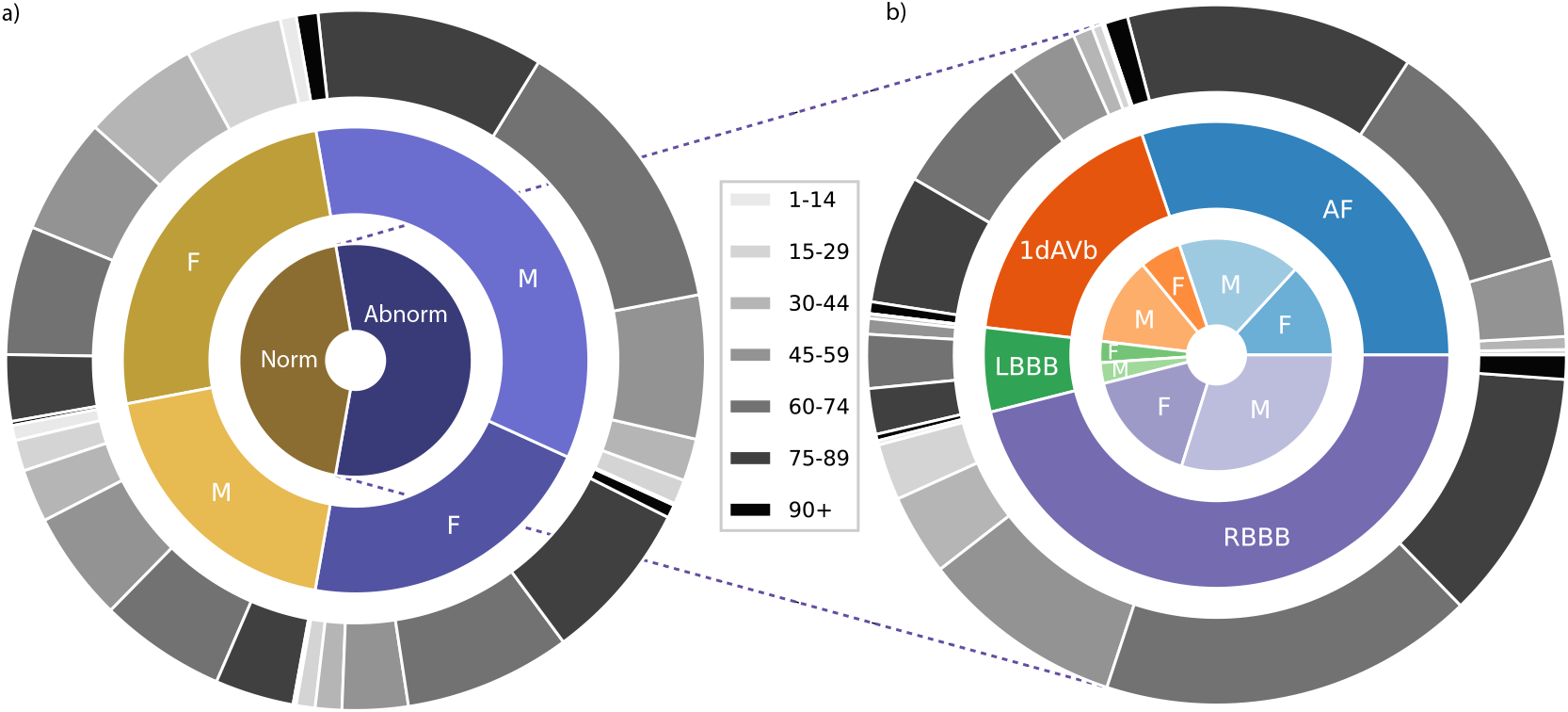
Distribution of patients with studied abnormalities versus those without abnormalities or with normal ECG readings in the CPSC dataset, along with a breakdown of specific individual abnormalities.

The analysis of the CPSC dataset revealed a gender disparity with a higher representation of male patients compared to female patients. However, the age distribution aligns with the general population, showing a more significant proportion of older individuals. Additionally, while most abnormalities are well represented in the dataset, a slight imbalance is observed in the occurrence of LBBB compared to other abnormalities.

## 4 Methods

To assess the viability of our proposed technique, we’ve developed a comprehensive experiment methodology detailed in Fig. 2. The entire process adheres to the steps illustrated in the figure.

The entire process can be segmented into three significant steps, each of which will be elaborated upon in the following subsections.

### 4.1 Step 1 - Training of *µ*-Trainer

The initial step of the entire process is conducted externally to the device and involves creating a compact model that mirrors the behavior of the larger model. This smaller model will be utilized in subsequent steps for on-device fine-tuning using personal data. This approach enables on-device fine-tuning since the larger model demands significant computational resources for training, rendering it impractical for on-device implementation. As a result, the larger model is exclusively used for inference on-device, while its smaller counterpart is employed for fine-tuning personal data.

The initial model utilized in this experiment is derived from Ribeiro et al. [14]’s research, which was designed for abnormality detection using 12-lead ECG data. A reduced version of the original model termed the “Tiny Trainer,” was developed for this study. Specifically, the third and fourth layers of the original model were omitted to create the *µ*-Trainer. As a result, while the *µ*-Trainer shares a similar architectural framework with the Original Model, it diverges regarding layer count and weight distribution.

The training began by initializing the weights of the first and bottom layers of the *µ*-Trainer with those from the corresponding layers of the original model. Subsequently, the weights of the top and bottom layers of the *µ*-Trainer remained constant throughout the training phase. To achieve this, a method called KD was used to train the *µ*-Trainer model to mimic the original model’s behavior closely. The main goal of this approach was to ensure a high degree of alignment in the behavior of the middle layers between the Original Model and the *mu* trainer. The entire training process was conducted using the TNMG subset, as the pre-trained Original Model had initially been trained on the TNMG dataset.

### 4.2 Step 2 - On-Device Fine-tuning

The second step involves the fine-tuning process, intended to be carried out on-device, allowing the model to be personalized using individual data for personal use. By conducting this process on-device, there is no need to transfer personal data externally. This approach safeguards user privacy and conserves device energy, as transmitting large amounts of sensitive data wirelessly consumes considerable (electrical) power, particularly in medical devices, which is crucial for the next generation of wearable and implantable systems.

During this fine-tuning process, the middle layers of the *µ*-Trainer are kept frozen throughout, ensuring that they retain the knowledge and patterns learned from the original training. Meanwhile, the top and bottom layers can adapt and learn from the personalized data, enabling the model to refine its predictions based on individualized information.

The CPSC dataset was employed to simulate the fine-tuning process on personal data, enabling the observation and analysis of performance changes before and after the fine-tuning procedure. This methodology facilitates a thorough assessment of how the model’s performance evolves and improves when fine-tuned using targeted datasets like CPSC.

### 4.3 Step 3 - Updating Weights on Original Model

In this step, the main goal is to use the fine-tuned *µ*-Trainer to update the weights of the Original Model. This will result in an updated model that delivers enhanced inference results tailored to personalized usage. Depending on the subsequent action phase, the updating process can be executed either on-device or externally. During this step, the weights of the top and bottom layers of the Original Model are directly replaced with the corresponding layers of the fine-tuned *µ*-Trainer. This integration of personalized learning into the Updated Model ensures that it is optimized for future on-device inference for personal usage.

### 4.4 Naive Continual Fine-tuning Process

After detailing the entire process, a continual fine-tuning process is proposed to handle incoming new data streams. This process is summarized in Fig. 3. The iterative nature of this continual learning process applies the presented methodology to learn from new data streams. The process is divided into two steps: the first step mirrors that of the previously explained method, while the second step combines the two steps proposed in the method. Subsequently, the Updated Model replaces the Original Model for the next iteration.

### 4.5 Evaluation Metrics

Evaluation metrics are crucial to helping with meaningful model performance comparisons. Precision (*P*) 8 and recall (*R*) 7 represent fundamental metrics for model evaluation, assessing predictive accuracy, and identification of actual positives, respectively. The F1-score (*F*_1_) 9 harmonizes precision and recall, balancing their trade-off.

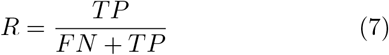

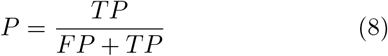

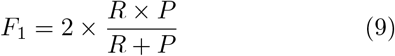

Area Under the Receiver Operating Characteristic Curve (AUROC) metric gauges a model’s ability to discriminate between negative and positive cases across various threshold levels, commonly applied in binary classification scenarios.

Area Under the Precision-Recall Curve (AUPRC), measuring the area under the precision-recall curve, offers critical insights into model performance, particularly in imbalanced binary classification tasks. Higher AUPRC scores signify a more balanced precision-recall relationship.

These evaluation metrics, including F1-score, precision, recall, AUROC, and AUPRC, are pivotal in assessing machine learning models, especially in anomaly detection within binary classification contexts.

## 5 Experiment

The experiment mirrors the overall process by dividing it into three distinct stages, breaking the entire procedure into three smaller sections aligned with each method step.

### 5.1 Step 1 - Training of *µ*-Trainer

In this process phase, we conducted experiments on several key aspects of the training process for the *µ*-Trainer model. Firstly, we focused on the model’s architecture, aiming to minimize the number of weights to ensure it remains lightweight enough for on-device fine-tuning. To achieve this, we initially tested incorporating a single layer of ResBlk between the top and bottom layers. However, despite significantly reducing the number of weights, this led to a considerable decline in the model’s performance.

Secondly, we explored using KD during the training process of the *µ*-Trainer. Although it’s possible to train the model without KD, we observed that it does impact the model’s performance in subsequent stages.

Lastly, we delved into the choice of loss function for KD, which is a critical factor to consider. Given that the model is tailored for multi-labeled classification, where individuals may have multiple abnormalities simultaneously, determining an appropriate loss function for distilling a multi-labeled model remains an area of ongoing exploration. In this experiment, we made initial forays into this domain to identify a suitable loss function from existing methodologies.

The Kullback-Leibler (KL) divergence is a widely used loss function in KD for classification models. However, based on experiments, its efficiency in distilling multi-labeled models is limited. Therefore, we devised a custom loss function based on binary cross-entropy, which is well-suited for training multi-labeled models.

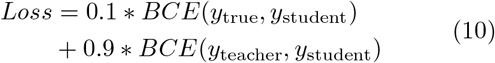

Equ. 10 illustrates the loss function employed in distilling the *µ*-Trainer model, with *BCE* denoting the Binary Cross-Entropy Loss function. Here, *y*_true_ denotes the actual labels assigned to each datum, while *y*_teacher_ signifies the binary predictions generated by the teacher model (specifically, the Original Model). On the other hand, *y*_student_ represents the probabilistic predictions generated by the student model (in this context, referring to the *µ*-Trainer).

The design of the loss function aims to align the behavior of the *µ*-Trainer model as closely as possible with that of the Original Model while ensuring a reasonable connection to the ground truth (i.e., the true labels).

### 5.2 Step 2 - Fine-tuning Process

In the process of fine-tuning, we aim to adjust the *µ*- Trainer model using the unfamiliar CPSC dataset. We conducted a series of evaluations on various aspects of this fine-tuning procedure. The CPSC dataset encompasses eight distinct classes of abnormalities, of which only four align with those found in the TNMG dataset. Consequently, our fine-tuning efforts are focused exclusively on these four shared classes.

To facilitate this fine-tuning, we employed a Binary Cross-Entropy loss function that was customized explicitly. This tailored loss function exclusively considers predictions related to those above four common classes while disregarding predictions related to the remaining classes. Batch size also plays a pivotal role in this context, with smaller batch sizes demanding less computational power and proving feasible for on-device training. However, excessively small batch sizes can impact the performance of the fine-tuned model.

The utilization of the Raspberry Pi 3 Model B Rev 1.2 to experiment with the on-device fine-tuning capability of the *µ*-Trainer is noteworthy. This device features a Broadcom BCM2387 chipset with a 1.2GHz Quad-Core ARM Cortex-A53 processor and 1GB of RAM. These specifications render the Raspberry Pi an optimal platform for conducting experiments and assessing the performance of the *µ*-Trainer, especially in resource-constrained environments.

The characteristics of the fine-tuning data also play a crucial role in fine-tuning the *µ*-Trainer model. Since the CPSC dataset is highly unbalanced, a smaller but balanced dataset was extracted from CPSC to investigate whether the behavior of the fine-tuned model would be affected.

### 5.3 Step 3 - Weights Update

Updating the weights from the fine-tuned *µ*-Trainer to the Original Model facilitates the ultimate comparison between the Updated and Original Models. This comparison is primarily carried out on a separate test set derived from the CPSC dataset, which was not utilized during the fine-tuning process.

Furthermore, we can also analyze the behavioral differences between the models on the dataset they were initially trained on (TNMG dataset).

### 5.4 Naive Continual Fine-tuning Process

After completing the experiments throughout the overall process, we propose a continuous fine-tuning approach to fine-tune the model privately when new data streams are collected from the device.

This experiment provides an initial demonstration of the feasibility of such a mechanism. Due to limited data availability, the experiment involved only a few iterations, each on a subset of the dataset.

## 6 Result

This section will present the results obtained from the designed experiments in the order presented in the previous section.

### 6.1 Training of *µ*-Trainer

During the initial stages of the experiment, we faced the primary challenge of integrating KD into the training process. Researchers have relatively understudied the application of a KD loss function for a Multi-Label model. Despite testing widely used loss functions for KD, such as KL divergence, we encountered unsatisfactory training results. In particular, the *µ*-Trainer did not effectively acquire knowledge from the Original Model. Consequently, we opted to utilize the proposed loss function for KD in training the *µ*-Trainer.

Given the preference for a smaller *µ*-Trainer for on-device fine-tuning, we reduced the size of the Original Model by removing several layers. Initially, we tested a *µ*-Trainer with just one ResBlk layer to assess its performance compared to the Original Model. This *µ*-Trainer was trained using the TNMG subset, the dataset used to create the Original Model. We then evaluated and compared the performance of the *µ*-Trainer with the Original Model on a test set within the subset. The results are presented in the upper part of Table 3, providing a summary of the Original Model’s performance and displaying the performance of the *µ*-Trainer with a single ResBlk layer trained using KD, tested on the test set.

**Table 3:** Models Tested on TNMG (P: Precision, R: Recall)

| Class | P | R | F1 | AUROC | AUPRC | P | R | F1 | AUROC | AUPRC |
| --- | --- | --- | --- | --- | --- | --- | --- | --- | --- | --- |
| Original Model | | | | | | 1-ResBlk μ-Trainer w/ KD | | | | |
| 1dAVb | 89.98% | 63.81% | 74.67% | 97.34% | 88.54% | 35.29% | 1.94% | 3.68% | 68.61% | 25.60% |
| RBBB | 97.51% | 90.53% | 93.89% | 99.39% | 98.24% | 92.25% | 86.61% | 89.34% | 97.04% | 92.86% |
| LBBB | 96.47% | 88.01% | 92.05% | 99.59% | 98.13% | 94.70% | 83.63% | 88.82% | 98.31% | 95.21% |
| SB | 95.00% | 79.78% | 86.73% | 99.40% | 96.07% | 79.02% | 52.72% | 63.25% | 93.24% | 71.63% |
| AF | 98.00% | 81.67% | 89.09% | 98.67% | 96.58% | 80.04% | 55.69% | 65.68% | 89.69% | 73.33% |
| ST | 96.86% | 85.67% | 90.92% | 99.66% | 98.30% | 83.94% | 70.88% | 76.86% | 97.14% | 86.64% |
| Average | 95.64% | 81.58% | 87.89% | 99.01% | 95.98% | 77.54% | 58.58% | 64.60% | 90.67% | 74.21% |
| 2-ResBlk μ-Trainer w/ KD | | | | | | 2-ResBlk μ-Trainer w/o KD | | | | |
| 1dAVb | 81.76% | 60.10% | 69.27% | 93.60% | 78.42% | 69.63% | 84.81% | 76.47% | 95.97% | 83.86% |
| RBBB | 97.84% | 89.03% | 93.23% | 98.58% | 96.87% | 95.25% | 90.30% | 92.71% | 98.46% | 96.36% |
| LBBB | 95.13% | 88.60% | 91.75% | 99.24% | 97.17% | 84.66% | 96.05% | 90.00% | 99.21% | 96.56% |
| SB | 85.95% | 81.80% | 83.82% | 98.09% | 89.77% | 66.10% | 96.11% | 78.33% | 97.92% | 90.30% |
| AF | 94.13% | 77.92% | 85.26% | 97.24% | 92.86% | 87.65% | 83.75% | 85.65% | 97.04% | 91.50% |
| ST | 92.01% | 85.21% | 88.48% | 98.62% | 94.97% | 90.07% | 85.21% | 87.57% | 99.01% | 94.69% |
| Average | 91.14% | 80.44% | 85.30% | 97.56% | 91.68% | 82.23% | 89.37% | 85.12% | 97.93% | 92.21% |

Compared to the Original Model, the *µ*-Trainer with 1 ResBlk layer performs significantly differently, especially for the 1dAVb condition, where a substantial performance gap is observed. This indicates that the *µ*-Trainer fails to mimic the performance of the Original Model, making it an unsuitable candidate for the fine-tuning process. A good trainer should replicate the performance of the Original Model as closely as possible.

We expanded the ResBlk layers to two and employed distinct training strategies: one with KD and the other without. The ensuing performance comparison is summarized in the lower part of Table 3, both with the KD strategy and without the KD strategy.

While both strategies produce effective trainers, achieving relatively high performance in the test set, there is an observable difference. The *µ*-Trainer trained using KD exhibits performance closer to the Original Model, as both have higher precision than recall. In contrast, the *µ*-Trainer trained without KD shows higher recall than precision. This indicates that the KD-trained trainer behaves more similarly to the Original Model and is thus a better candidate for the fine-tuning process.

The *µ*-Trainer trained with KD was employed as the primary model from this stage onward. As a baseline comparison, the Original Model was tested on the CPSC test set to evaluate its performance without fine-tuning. Only the four abnormalities common with TNMG were considered. The inference results are summarized in Table 4.

**Table 4:**
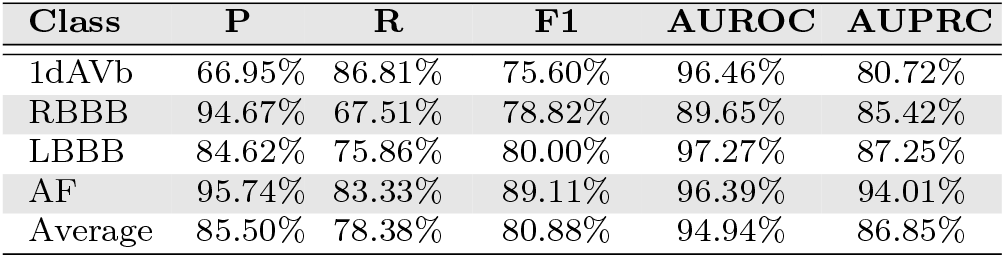
Testing the Original Model on CPSC Test Set (P: Precision, R: Recall)

As previously mentioned, the dataset’s characteristics used for fine-tuning are crucial. In our experiment, we fine-tuned the entire available CPSC dataset, which comprises 6000 data points used for fine-tuning. Additionally, we created a smaller balanced dataset from the CPSC containing 200 instances of each abnormality and 200 instances without any of the four abnormalities. The results are presented in the upper part of Table 5.

**Table 5:** Fine-Tuned Models Testing on CPSC Test Set (P: Precision, R: Recall)

| Class | P | R | F1 | AUROC | AUPRC | P | R | F1 | AUROC | AUPRC |
| --- | --- | --- | --- | --- | --- | --- | --- | --- | --- | --- |
| **Whole-Dataset Fine-Tuned** |  |  |  |  |  | **Balanced-Dataset Fine-Tuned** |  |  |  |  |
| 1dAVb | 58.74% | 92.31% | 71.79% | 96.57% | 76.46% | 66.13% | 90.11% | 76.28% | 96.55% | 80.51% |
| RBBB | 92.93% | 72.15% | 81.24% | 93.76% | 90.75% | 94.77% | 68.78% | 79.71% | 90.58% | 86.56% |
| LBBB | 92.31% | 82.76% | 87.27% | 99.58% | 92.33% | 84.62% | 75.86% | 80.00% | 97.89% | 87.88% |
| AF | 96.90% | 77.16% | 85.91% | 96.36% | 94.36% | 95.74% | 83.33% | 89.11% | 96.42% | 94.20% |
| Average | 85.22% | 81.09% | 81.55% | 96.57% | 88.48% | 85.31% | 79.52% | 81.27% | 95.36% | 87.29% |
| **Whole-Dataset On-Device Fine-Tuned** |  |  |  |  |  | **Balanced-Dataset On-Device Fine-Tuned** |  |  |  |  |
| 1dAVb | 58.74% | 92.31% | 71.79% | 96.50% | 76.30% | 58.74% | 92.31% | 71.79% | 96.43% | 75.80% |
| RBBB | 94.89% | 70.46% | 80.87% | 93.56% | 90.50% | 93.44% | 72.15% | 81.43% | 93.81% | 90.77% |
| LBBB | 88.89% | 82.76% | 85.71% | 99.53% | 91.61% | 88.89% | 82.76% | 85.71% | 99.57% | 92.62% |
| AF | 97.62% | 75.93% | 85.42% | 96.35% | 94.39% | 96.88% | 76.54% | 85.52% | 96.23% | 94.12% |
| Average | 85.03% | 80.36% | 80.95% | 96.48% | 88.20% | 84.49% | 80.94% | 81.11% | 96.51% | 88.33% |

While the fine-tuned model on the entire CPSC dataset shows a larger average improvement overall, the model fine-tuned on the balanced subset demonstrates a more balanced improvement across various metrics. Notably, an improvement is observed in almost every metric. It’s worth mentioning that the balanced dataset comprises only 1000 instances, which is one-sixth of the whole dataset.

We then replicated this process on a Raspberry Pi 3 Model B Rev 1.2. The entire fine-tuning process was completed, albeit with limited memory. The batch size was reduced from 32 to 8 to ensure successful fine-tuning on the device. Approximately 70-80% of the memory was utilized during the fine-tuning process. The results for fine-tuning the entire dataset and the balanced subset are summarized in the lower part of Table 5.

The decrease in batch size impacts the performance of both fine-tuned models trained on datasets of varying sizes. However, the model trained on the balanced dataset performs slightly better than the others. Although we observe overall improvements in both fine-tuned models, the model trained on the balanced set performs slightly better. Notably, the 1dAVb class is the most affected, possibly because the Original Model exhibits the poorest performance among the four classes we are evaluating. This highlights the model’s somewhat limited capability for detecting the 1dAVb condition.

The fine-tuned models improved the fine-tuned dataset, demonstrating their adaptability to data using the proposed methodology. Subsequently, we evaluated the performance of the fine-tuned model on the TNMG dataset, which was the model’s original training dataset. This assessment provides insights into the fine-tuned model’s behavior following the fine-tuning process on the original dataset. Table 6 presents a summary of the model’s performance after undergoing fine-tuning with either the complete CPSC dataset or a balanced CPSC subset on the Raspberry Pi 3. This evaluation was conducted using the TNMG test set.

**Table 6:** On-Device Fine-Tuned Models Tested on TNMG Test Set (P: Precision, R: Recall)

| Class | P | R | F1 | AUROC | AUPRC | P | R | F1 | AUROC | AUPRC |
| --- | --- | --- | --- | --- | --- | --- | --- | --- | --- | --- |
| With Full CPSC Data |  |  |  |  |  | With CPSC Balanced Subset |  |  |  |  |
| 1dAVb | 94.30% | 29.40% | 44.83% | 89.52% | 77.64% | 93.24% | 31.18% | 46.73% | 91.26% | 79.38% |
| RBBB | 98.26% | 71.82% | 82.99% | 95.51% | 93.56% | 97.76% | 75.75% | 85.36% | 96.38% | 94.37% |
| LBBB | 97.83% | 52.63% | 68.44% | 93.55% | 88.71% | 97.99% | 57.02% | 72.09% | 94.11% | 89.45% |
| SB | 96.25% | 47.90% | 63.97% | 93.27% | 86.81% | 96.96% | 49.61% | 65.64% | 94.08% | 87.90% |
| AF | 99.66% | 41.11% | 58.21% | 91.39% | 86.54% | 99.70% | 45.56% | 62.54% | 92.78% | 88.24% |
| ST | 98.41% | 57.32% | 72.44% | 95.87% | 91.93% | 98.25% | 60.55% | 74.93% | 96.74% | 92.99% |
| Average | 97.45% | 50.03% | 65.15% | 93.19% | 87.53% | 97.32% | 53.28% | 67.88% | 94.22% | 88.72% |
| With Full CPSC Data (Threshold 20%) |  |  |  |  |  | With CPSC Balanced Subset (Threshold 20%) |  |  |  |  |
| 1dAVb | 84.23% | 63.00% | 72.09% | 89.52% | 77.64% | 84.21% | 64.62% | 73.13% | 91.26% | 79.38% |
| RBBB | 93.69% | 83.95% | 88.55% | 95.51% | 93.56% | 93.63% | 84.87% | 89.04% | 96.38% | 94.37% |
| LBBB | 93.60% | 74.85% | 83.18% | 93.55% | 88.71% | 93.89% | 74.12% | 82.84% | 94.11% | 89.45% |
| SB | 92.93% | 71.54% | 80.84% | 93.27% | 86.81% | 93.28% | 73.41% | 82.16% | 94.08% | 87.90% |
| AF | 95.68% | 64.58% | 77.11% | 91.39% | 86.54% | 96.50% | 65.14% | 77.78% | 92.78% | 88.24% |
| ST | 94.07% | 78.27% | 85.45% | 95.87% | 91.93% | 93.68% | 79.97% | 86.28% | 96.74% | 92.99% |
| Average | 92.37% | 72.70% | 81.21% | 93.19% | 87.53% | 92.53% | 73.69% | 81.87% | 94.22% | 88.72% |

While both cases exhibit very high AUROC and AUPRC performance, the recall of the fine-tuned models is relatively low. However, when the threshold is reduced from the original 50% to 20%, the performances for fine-tuning with the full CPSC dataset and the balanced CPSC subset are displayed in the lower part of Table 6. The recall is now higher than previously observed while maintaining a high precision reading. This indicates that after the fine-tuning process, the models remain generalizable to the original data used to train the Original Model, although they are less confident as the probabilities they predict are slightly lower.

Comparing the performance between the models trained on the full CPSC dataset and the balanced CPSC subset, the model fine-tuned on the CPSC balanced subset slightly outperforms the former. Considering that the amount of data in the balanced CPSC subset is only one-sixth of the complete CPSC data used in fine-tuning, a balanced dataset enhances the model’s generalizability.

### 6.4 Naive Continual Fine-tuning Process

To establish a scenario involving the reception of new data streams and the application of our proposed continuous fine-tuning process to a limited dataset, we partitioned the CPSC dataset into two subsets, each comprising 3,000 data points. Each subset was subsequently employed in one iteration to evaluate the effectiveness of our proposed method. It is crucial to mention that this process was not conducted on the edge, as this preliminary study was focused on assessing the efficacy of the proposed continual fine-tuning process. The comprehensive results of the iterative analysis are outlined in Table 7.

**Table 7:** Average Performance of Proposed Continual Learning Process (P: Precision, R: Recall)

| Iteration | P | R | F1 | AUROC | AUPRC |
| --- | --- | --- | --- | --- | --- |
| Zero | 85.50% | 78.38% | 80.88% | 94.94% | 86.85% |
| One | 84.94% | 79.16% | 80.87% | 95.90% | 87.70% |
| Two | 85.31% | 79.95% | 81.06% | 96.56% | 88.75% |

As the number of iterations increases, it becomes evident that the performance of the fine-tuned Updated Model improves. This enhancement is notably prominent in the AUROC and AUPRC scores, which assess the distinction between positive/negative and precision/recall. While precision, recall, and F1 score can be adjusted by threshold, AUROC and AUPRC demonstrate the model’s growing confidence with each iteration.

## 7 Discussion

The Original Model used to demonstrate the case is a multi-label model, showcasing that the fine-tuning process can have varying impacts on the abnormalities we aim to detect. It has been observed that the improvement is stable and noticeable for specific abnormalities, whereas for others, the improvement is comparatively smaller. This observation may be linked to the Original Model performing better on some abnormalities while performing less effectively on others. In general, it has been observed that when the Original Model excels in a specific task, the fine-tuning process also yields more favorable results.

While our proposed method has demonstrated improved performance through fine-tuning, the extent of improvement has been moderate rather than dramatic. This can be attributed to the fine-tuning process primarily focusing on the top and bottom layers of the model, which have a limited number of weights. Specifically, these layers comprise only 0.7% of the 6,425,638 total weights in the Original Model and 1.6% in the *µ*-Trainer, a scaled-down version of the Original Model. Increasing the weights of the top or bottom layers within a model has the potential to enhance performance, provided that due regard is given to the computational power and memory constraints inherent to the edged device. The efficacy of this strategy is closely intertwined with the model’s architectural design and the computational capabilities of the specific edged device in use.

The setup for this experiment is appropriate, especially considering the use of a relatively less powerful device like the Raspberry Pi 3, which can only handle on-device training for limited weights. Fine-tuning the Original Model on the device proved challenging despite attempting to freeze all layers except for the top and bottom ones. However, optimizing the model by increasing the weight density in the top and bottom layers in future iterations or real-world applications with more advanced hardware should lead to more substantial performance improvements. It’s essential to balance the number of trainable weights and the edge device’s power consumption to ensure efficient and practical training.

Our observations during the process have highlighted the significant impact of the *µ*-Trainer on the overall fine-tuning performance. We found that the closer the *µ*-Trainer’s behavior aligns with the Original Model, the more favorable the fine-tuning results tend to be. Conversely, deviations in behavior between the *µ*-Trainer and the Original Model lead to a noticeable decrease in fine-tuning performance. If the *µ*-Trainer exhibits a substantial performance disparity compared to the Original Model, this method may not be viable. Consequently, during the model design phase, it is imperative to ensure that the performance of the *µ*-Trainer closely aligns with that of the original model.

In addition to the *µ*-Trainer, we discovered that other hyper-parameters, such as batch size, also play a crucial role in determining the performance of our proposed method. Due to limited RAM in the devices, we had to use a batch size of 8 for on-device fine-tuning, which unfortunately resulted in a decrease in the overall performance of the proposed method.

It’s important to acknowledge that our proposed naive continual learning process is still in its preliminary stages. While we believe it holds promise for integration into a long-term fine-tuning personalization process, we used only two (2) subsets in the iterative fine-tuning process due to limited data availability. Despite demonstrating favorable results, we acknowledge ample room for further refinement and improvement of this method.

### 7.1 Privacy Protection

The proposed method, although preliminary, sheds light on addressing privacy issues. This method eliminates the need for data transmission from local devices to the cloud, which is the conventional approach, thus alleviating significant privacy concerns. Additionally, it tackles the challenges of FL, as previous research has indicated potential data replication through the gradients shared in the FL process [12] [13].

Moreover, FL needs to address the capacity constraints of local training, particularly in scenarios involving devices with comparatively lower computational capabilities. In contrast, the proposed method simultaneously addresses the local training capacity and privacy protection issues, making it a promising solution in this domain.

Applying the proposed method, the model update process is streamlined, avoiding iterative gradient updates typical in FL. Instead, only segments of the trained weights, generated after the fine-tuning process, are shared beyond the device. This strategic approach effectively mitigates data leakage risks, a contribution highlighted in various research papers [12] [13].

## 8 Limitations and Future Works

This study employs a multi-label model as its foundational framework. It is important to note that the knowledge distillation loss function, specifically in the context of multi-label models, remains an area that has yet to be explored. The choice of loss function significantly impacts the characteristics of the *µ*-Trainer, subsequently influencing the performance of the fine-tuning process. Through further research and exploration into the intricacies of the loss function, there is potential to enhance the overall performance of the proposed method.

The architecture design of the model was adapted from Ribeiro et al. [14]’s work rather than being specifically designed for the proposed methodology. As mentioned previously, the weights in the top and bottom layers of the model constitute only a small fraction of the total number of weights, thereby limiting the capacity for fine-tuning. It is crucial to consider the design of the architecture in conjunction with the device’s computational power. A balanced arrangement would yield optimal results.

The proposed Naive Continual Fine-tuning Process is currently at its preliminary stage, serving as a vision for future development. Due to the limited amount of streamed data utilized at this stage, there is a possibility that both the process and methodology may not be optimized, leaving ample room for future research and refinement.

## 9 Conclusion

This paper introduces a cutting-edge approach that leverages a smaller trainer and advanced knowledge distillation techniques to fine-tune a larger model to detect abnormalities. This model leverages a smaller trainer to perform on-device fine-tuning using personalized data, thereby facilitating efficient on-device inference. The results of fine-tuning the larger model indicate a notable improvement, achieving an average AURPC of 96.51% compared to the pre-fine-tuning result of 94.94%. The impact of fine-tuning varies across different abnormalities, indicating that the extent of improvement relies on the Original Model’s performance in specific tasks. Notably, ensuring alignment between the behavior of the *µ*-Trainer and the Original Model results in more favorable fine-tuning outcomes, underscoring the significance of maintaining consistency in model behavior during the fine-tuning process. While this method demonstrates promising potential in enhancing model performance through fine-tuning, further refinement and exploration is crucial to fully unlock its capabilities, especially in the proposed Continual Fine-tuning Process. This research lays the groundwork for future advancements in personalized model adaptation and more precise on-device inference and emphasizes the importance of protecting user data privacy and reducing the cost of transmitting data throughout these developments.

## Data Availability

This research paper uses the public CPSC dataset but acknowledges that the TNMG dataset is private and requires permission from its owner for access.

## 10 Availability of Code

You can access the code through this link: https://github.com/NeuroSyd/microTrainer. Please be aware that specific terms, conditions, or usage restrictions may apply to the code.

## Notes

### Competing Interest Statement

The authors have declared no competing interest.

### Funding Statement

This study did not receive any funding

